# Rice Diet Improves Survival, Blood Pressure, and Eye Changes in 544 Patients with Malignant-Hypertension

**DOI:** 10.64898/2026.01.09.26343821

**Authors:** Francis A. Neelon, Philip J. Klemmer, Jong Ok La, Scott L. Sanoff, Yi-Ju Li, Anthony N. Kuo, Crystal C. Tyson, Anastacia Bohannon, William McDowell, Fredrich C. Luft, Pao-Hwa Lin

**Affiliations:** Department of Medicine, Duke University School of Medicine, Durham, NC; School of Medicine, University of North Carolina at Chapel Hill, Chapel Hill, NC; Duke Molecular Physiology Institute, Duke University School of Medicine, Durham, NC; Division of Nephrology, Department of Medicine, Duke University School of Medicine, Durham, NC; Department of Biostatistics and Bioinformatics, Duke University School of Medicine, Durham, NC; Department of Ophthalmology, Duke University School of Medicine, Durham, NC; Experimental and Clinical Research Center (ECRC), a cooperation of Charité - Universitätsmedizin Berlin and Max Delbrück Center for Molecular Medicine (MDC), Berlin, Germany

**Keywords:** malignant hypertension, low sodium, rice diet, survival, retinal hemorrhage, papilledema

## Abstract

**Background:** The sodium-restricted rice diet (RD) was once the only effective treatment for malignant hypertension (MH); however, a modern, comprehensive data analysis is lacking. We determined patient survival and ocular improvements in 544 MH patients treated between 1942-1982.

**Methods:** At entry, systolic blood pressure (SBP) was ≥170 mmHg and retinal hemorrhage (n = 312), hemorrhage with papilledema (n= 211) or papilledema alone (n = 21) were present. Dates of death were available for 454 patients; ocular data (at baseline and again before day 365) for 342 patients with hemorrhage and 143 with papilledema. We used actuarial analysis to determine survival and resolution of ocular findings. We used Cox proportional hazards to calculate mortality hazard ratio (HR), and period life tables to estimate loss of longevity.

**Results:** Median initial SBP of 213.3 mmHg fell to 178.4 during year 1, and to ∼143 after 9 years. RD patients survived longer than untreated patients: 1890 vs 540 days for patients with hemorrhage alone; 510 vs 180 days with both hemorrhage and papilledema. Few patients reached their expected longevity; median loss of potential life was 15.4 years. Compared to patients whose SBP fell <15 mmHg by 4 weeks, those with a fall ≥ 37 mmHg had HR for mortality of 0.32. Retinal hemorrhages cleared in 260/342 patients; papilledema, in 133/143.

**Conclusion:** With RD treatment blood pressure decreased, and ocular abnormalities largely resolved. Survival improved, but predicted longevity was not achieved. The RD helped MH and could still provide a useful adjunct to pharmacologic therapy.

## BACKGROUND

The hallmark of malignant hypertension (MH), the very thing that creates its malignancy, is the profound truncation of life span imposed on those affected. In 1939, Keith, Wagener and Barker (KWB)^1^ recorded a median survival after diagnosis of ∼520 days in 37 untreated patients with severe hypertension accompanied by retinal hemorrhages (H), and of ∼160 days in those with both papilledema (P) and H. There was no satisfactory or truly effective treatment until about 1940, when Walter Kempner began treating MH patients with his low-sodium rice diet (RD).

In 1955, Newborg and Kempner reported that, in 120 patients with H and P treated with the RD, patients with severely impaired kidney function had shorter survival, and those who adhered better to the diet, longer^2^; these patients are included in the present cohort (see Sanoff, et al for details on cohort assembly^3^). This report analyzes the long-term effects on blood pressure (BP), patient survival, and ocular changes of MH patients treated with the RD.

## METHODS

All patients were evaluated at Duke Hospital in Durham, North Carolina; they were then in residence in Durham for varying periods, eating at dedicated facilities adhering to the strictures of the RD; patients then returned home elsewhere, often coming back to Durham at intervals for evaluation and dietary re-emphasis. The RD was composed of rice and fruit, sometimes supplemented with vegetables and, occasionally, meat; it delivered <150 mg/day of sodium (see Sommerfeld, et al^4^ for details). Because direct measurement of urinary sodium was not available, Kempner used twice weekly measurements of chloride concentration in 24-hour urine collections to monitor dietary compliance.

### Patient Data

We established an electronic RD project database by digitizing hand-written paper records, maintained in the Duke University Medical Archives, of more than 17,000 patients treated with the RD.^4^ BP was measured by an experienced examiner using a mercury or aneroid sphygmomanometer, at about the same time each day, with the patient recumbent and after resting for ∼20 minutes. We identified 807 patients with systolic blood pressure (SBP) ≥170 mmHg recorded at least once between 7 days before and 6 days after starting the RD and with concomitant retinal H, P, or both recorded within 30 days before or after starting the RD; 232 were excluded because of coexisting conditions that might cause misclassification (diabetes mellitus, brain tumor) or misinterpretation of response (surgical sympathectomy), and another 31 because they were treated with the RD for <7 days.^3^ We defined baseline BP as the average of all BP recorded between days -7 to +1 of starting the RD. The 544 patients, aged 19-76 years, with severe hypertension and the ocular fundus abnormalities of MH, documented by retinal photography, form the basis of this report.

Following the terminology of KWB, we grouped patients with MH according to ocular findings: those with H but without P are designated as having Group III hypertension; those with both H and P as having Group IV; we have added a third group, termed Group V (P without H), a group not recorded by KWB^1^ but described by Lip et al^5^.

Because Kempner saw clinical improvement with RD treatment, he did not follow untreated MH patients but instead treated everyone who came to his clinic; we, therefore, compare survival of RD patients to the near-contemporary cohort described by KWB.^1^

The Duke University Medical Center Institutional Review Board (Pro00105257) approved construction of the database and execution of the study. Data can be accessed in the Duke Research Data Repository at https://research.repository.duke.edu/.

### Excess Blood Pressure

We calculated BPs in excess of desirable values (120 mmHg systolic or 80 mmHg diastolic) by subtracting 120 from each available SBP value and 80 from each available DBP. We then expressed the resulting differences as percentage of initial excess value (e.g., [Excess SBP/Initial Excess SBP]*100).

### Survival Outcome

To determine survival after treatment with the RD, we used dates of death recorded in the patient record or available from public death records (total, 454 patients). Survival was calculated as days from beginning the RD treatment until death. For those without death records, survival was censored at time of loss to follow-up. For patients whose date of death was only approximated (e.g., “died June, 1963” or “died 1954”), we used the mid-point of the possible time interval (i.e., “June 15, 1963” or “July 1, 1954”).

### Resolution of Ocular Fundus Abnormalities

In addition to ocular fundus photographs taken within ±30 days of starting the RD, we have data from photographs taken at least once more during the ensuing year on 342 patients with H (with or without P, 293 of whom completed at least one year of follow-up, and 49 of whom died or were lost to follow-up during that year), and on 143 patients with P (with or without H, 111 of whom completed at least one year of follow-up and 32 of whom died or were lost to follow-up). These data allow estimation of the time interval from start of RD treatment to resolution of retinal H and P.

A caveat regarding the ocular data is important here. Since the RD was a medical practice, not a protocol-driven experimental study, fundus photographs were taken at variable (sometimes many months-long) intervals. To accommodate this variability, we assumed that fundus changes persisted until the date of the first negative photographs, even though undocumented resolution might have occurred earlier.

### Predicted life-Span Analysis

To estimate predicted longevity, we used age-specific expected number of years of life derived from the period life-tables in the 2024 Trustees Report (TR2025) of the US Social Security Administration.^6^ Because period life tables are not available before 1900, we calculated, for years 1901 through1914, the ratio of expected years of life relative to the values in year 1900 for males and females aged 20, 30, 40, 50, 60, and 70. We averaged these sex- and age-specific ratios to derive yearly changes in life expectancy relative to year 1900, then fitted a least-squares mean trendline (scaled to equal 1.0 at year 1900) for years 1900 through 1914 and extrapolated backward to adjust expected years of life for patients born before 1900.

This correction decreased the calculated difference between median expected years of life expected and actual survival by only 0.283 years (2.1%). Years of potential life lost represent the difference between a patient’s actual survival and the survival predicted from the life-tables.

### Statistical Analysis

Descriptive statistics were computed for demographic and clinical characteristics for each MH group, summarized as frequency (percentage) for categorical variables, and median (quartiles Q1, Q3) for continuous variables. Pairwise differences in demographic and clinical characteristics for each hypertension group were compared using the t-test or Wilcoxon rank sum test for continuous variables, as appropriate, and Fisher’s exact test for categorical variables. Characteristics of patients who died before 90 days and those who survived beyond 120 days after starting the RD were compared using the same statistical methods as used for the full study population. SBP pattern for all MH patients available at yearly intervals up to 17 years after starting the RD was illustrated by a box plot of the medians, IQR, and total ranges of the time-interval average SBP for each individual.

Because the exact date of events of interest (death or resolution of ocular changes) is not known precisely for every patient, instead of Kaplan-Meier survival estimation, we used actuarial analysis^7^, which requires that the period of follow-up after starting treatment be divided into arbitrary but equally spaced epochs of time (we used 30-day intervals). To derive historical referents for untreated patients with Group III and IV hypertension, we calculated actuarial survival probabilities for Group III and Group IV, using data extracted from Figure 1 of KWB.^1^ Survival rates (time to death) after starting the RD were compared between MH groups, as was time to resolution for H and P.

**Figure 1.**
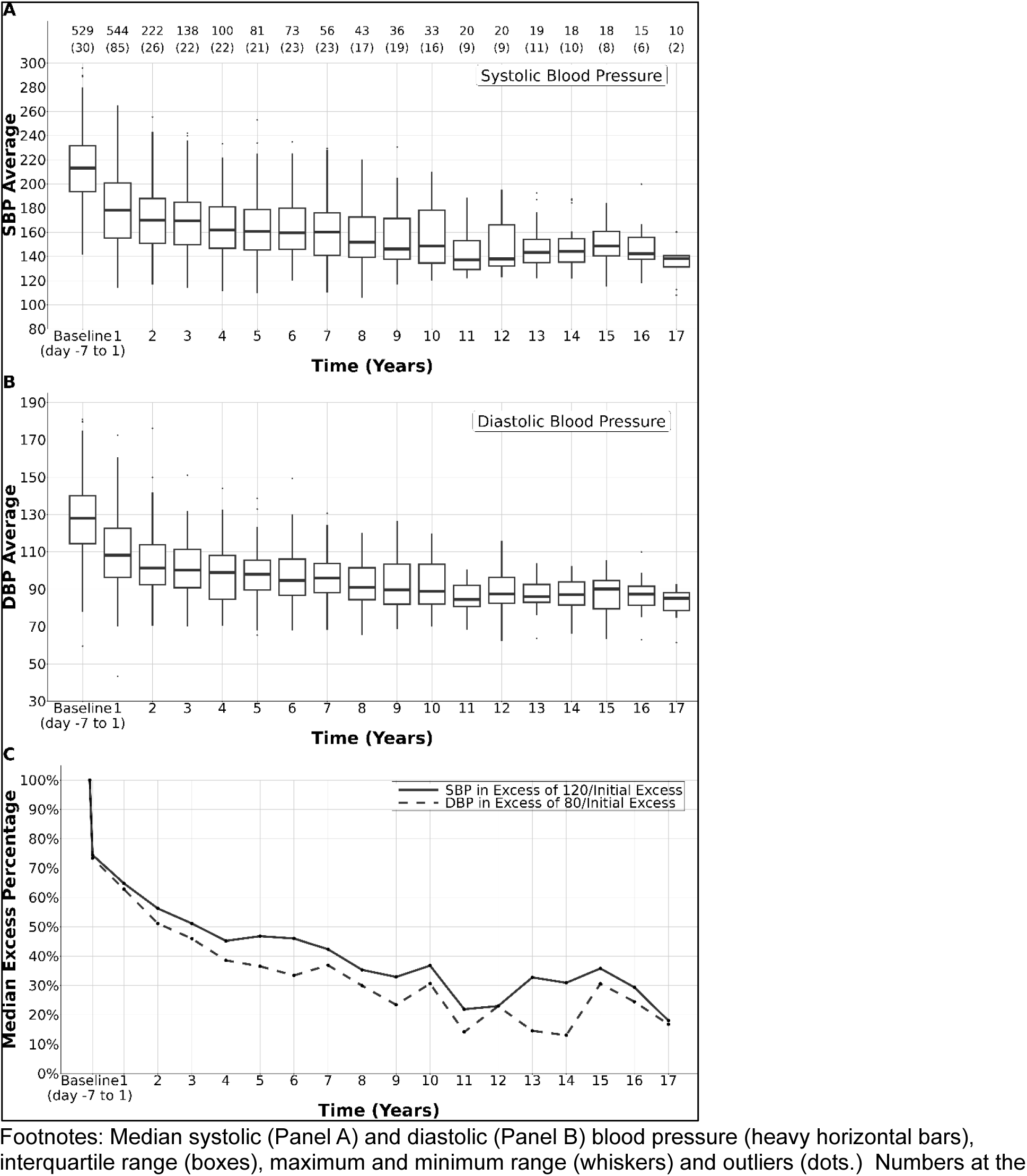
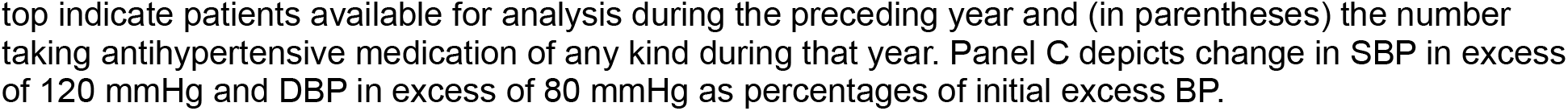
Blood pressure over 17 years after initial treatment with rice diet.

To investigate the relationship between long-term mortality and SBP change, we calculated the average change from baseline in available SBP over the first 4 weeks of RD treatment, categorized the change into quartiles, then used Cox proportional hazards modeling^8^ to test the effects of BP response on survival. We used four cycles of backward selection, eliminating the least significant variable at each step, to determine the final multivariable model. All analyses were performed using R.4.4.3. Significance was determined based on p <0.05.

## RESULTS

**Table 1** displays patient characteristics, by hypertension group, of the 544 MH patients: 312 had retinal H without P (Group III), 211 had H with P (Group IV) and 21 had P without H (Group V). Most patients (68.9%) were men; median age when starting RD was 54 years for Group III, 46 years for Group IV, and 42 years for Group V. Ages ranged from 19-76 years for the entire cohort. Median body mass index (BMI; weight in kg divided by height in m^2^) of the entire cohort was 23.5 kg/m^2^; BMI was <25 in 130, 25-29.9 in 53, and >30 in 18 patients. Unfortunately, missing height or weight data excluded 343 patients from BMI calculation at baseline and 374 at 4 weeks. Median systolic and diastolic (DBP) blood pressures were high at baseline (206/120 mm Hg in Group III; 223/137 in Group IV; 212/133 in Group V) and decreased by 21-37/14-19 mmHg after 4 weeks. Renal function was impaired at baseline as shown by median serum non-protein nitrogen (NPN) values of 38-51 mg/dL (reference range, 25-35 mg/dL) and blood urea nitrogen (BUN) of 16.8-27.4 mg/dL (reference range 7-15 mg/dL). After 4 weeks, those markers increased in both Group V and, especially, Group IV patients. Median baseline urine chloride concentration (UCl) was 217 mg/dl, consistent with a dietary sodium chloride intake of about 1500 mg/day, and fell by ∼90% after 4 weeks.

**Table 1.** Clinical characteristics of 544 MH patients, sorted by hypertension group*.

|  | Group III<br>Hypertension<br>n=312 |  | Group IV<br>Hypertension<br>n=211 |  | Group V<br>Hypertension<br>n=21 |  | P Value<sup>†</sup> |  |  |
| --- | --- | --- | --- | --- | --- | --- | --- | --- | --- |
|  | n<sup>‡</sup> |  | n<sup>‡</sup> |  | n<sup>‡</sup> |  | III vs IV | III vs V | IV vs V |
| Age, baseline, years | 312 | 54.0<br>[47.2, 59.1] | 211 | 46.3<br>[39.7, 54.0] | 21 | 41.8<br>[35.5, 46.4] | <0.001 | <0.001 | 0.083 |
| Male | 312 | 200<br>(64.1%) | 211 | 162<br>(76.8%) | 21 | 13 (61.9%) | 0.002 | 0.819 | 0.181 |
| BMI, baseline, kg/m<sup>2</sup> | 189 | 23.7<br>[21.3, 26.9] | 121 | 23.6<br>[21.5, 26.4] | 12 | 24.5<br>[20.4, 26.8] | 0.475 | 0.82 | 0.837 |
| BMI at wk4, kg/m<sup>2</sup> | 102 | 23.6<br>[21.2, 27.7] | 61 | 23.0<br>[19.4, 25.6] | 7 | 24.7<br>[21.6, 25.3] | 0.047 | 0.829 | 0.499 |
| SBP, baseline, mmHg | 303 | 207<br>[189, 223] | 208 | 223<br>[210, 238] | 19 | 212<br>[197, 246] | <0.001 | 0.28 | 0.304 |
| SBP at wk4, mmHg | 287 | 169<br>[149, 189] | 179 | 196<br>[175, 211] | 17 | 191<br>[141, 208] | <0.001 | 0.274 | 0.16 |
| DBP, baseline, mmHg | 303 | 120<br>[109, 134] | 208 | 137<br>[127, 146] | 19 | 133<br>[124, 142] | <0.001 | 0.005 | 0.327 |
| DBP, baseline, mmHg | 287 | 101<br>[91.7, 114] | 179 | 119<br>[106, 133] | 17 | 119<br>[101, 133] | <0.001 | 0.017 | 0.357 |
| NPN, baseline, mg/dL | 179 | 38.0<br>[33.5, 45.0] | 137 | 51.0<br>[39.0, 72.0] | 11 | 44.5<br>[37.0, 51.0] | <0.001 | 0.144 | 0.298 |
| NPN at wk4, mg/dL | 31 | 36.0<br>[32.5, 64.5] | 40 | 71.0<br>[50.4, 98.0] | 6 | 58.0<br>[38.8, 81.0] | 0.003 | 0.471 | 0.312 |
| BUN, baseline, mg/dL | 179 | 16.8<br>[13.7, 23.0] | 122 | 27.4<br>[17.9, 43.3] | 10 | 19.8<br>[16.5, 33.2] | <0.001 | 0.25 | 0.414 |
| BUN at wk4, mg/dL | 21 | 13.1<br>[9.50, 24.5] | 20 | 47.7<br>[30.7, 62.9] | 2 | 28.6<br>[18.2, 38.9] | 0.001 | 0.87 | 0.278 |
| UCL, baseline, mg/dL | 156 | 242<br>[117, 365] | 112 | 194<br>[108, 341] | 13 | 215<br>[102, 268] | 0.291 | 0.452 | 0.746 |
| UCL at wk4, mg/dL | 246 | 19.5<br>[13.0, 32.4] | 140 | 24.7<br>[15.5, 39.0] | 13 | 17.5<br>[14.3, 33.7] | 0.008 | 0.836 | 0.429 |

Footnotes: \*Median [Q1, Q3] or n (%). <sup>‡</sup>n=number available for analysis <sup>†</sup>t-test or Wilcoxon rank sum test for continuous variables, as appropriate, and Fisher's exact test for categorical variables. BMI= weight (kg) divided by height (m) squared; SBP=systolic blood pressure in mm Hg; DBP=diastolic blood pressure in mmHg; NPN=serum concentration of non-protein nitrogen in mg/dL; BUN=serum concentration of urea nitrogen in mg/dL; UCL=urine concentration of chloride in mg/dL.

### Long-term Effects on Blood Pressure

Panels A and B of **Figure 1** show median, interquartile and extreme ranges of interval-average SBP and DBP over 17 years after starting the RD, after which the paucity of patients precludes analysis. Of the 544 patients who qualified for inclusion in the study, 15 had no BP recorded within the specified baseline. During the first year, median SBP fell by 34.9 mmHg, from 213.3 to 178.4, stabilizing between years 9-17 at ∼143 mmHg; median DBP fell from 128 to 108.2 mmHg over the first year, reaching ∼87 mmHg after year 9. The number of patients available for analysis (shown in the line at the top of **Figure 1**) declined substantially over time, and the proportion taking anti-hypertensive medication (shown in parentheses above the figure) rose to ∼50% of patients by 9 years and thereafter.

Panel C of **Figure 1** shows that, during the first month on the RD, excesses of both SBP and DBP decreased by >30%, declining slowly thereafter and stabilizing at ∼25% of initial excess SBP and ∼15% of initial excess DBP between years 10-17.

### Survival after Rice Diet Treatment

Figure 2 shows estimates of actuarial survival for the 454 patients for whom we have dates of death. Dates of death were not available for 90 patients; compared to those with known dates of death, those without showed lesser male predominance, lesser SBP response at 4 weeks, lower DBP at baseline and after 4 weeks on the RD (supplemental Table S1). All differences were minimal and seem unlikely to alter conclusions about survival. Inset in Figure 2 is a depiction of the first 70 months of treatment, comparing RD patients in Groups III and IV to the untreated patients in those groups reported by KWB^1^ (who did not describe a Group V). The curves for RD patients closely track those of KWB patients until 90-120 days, after which they diverge.

**Figure 2.**
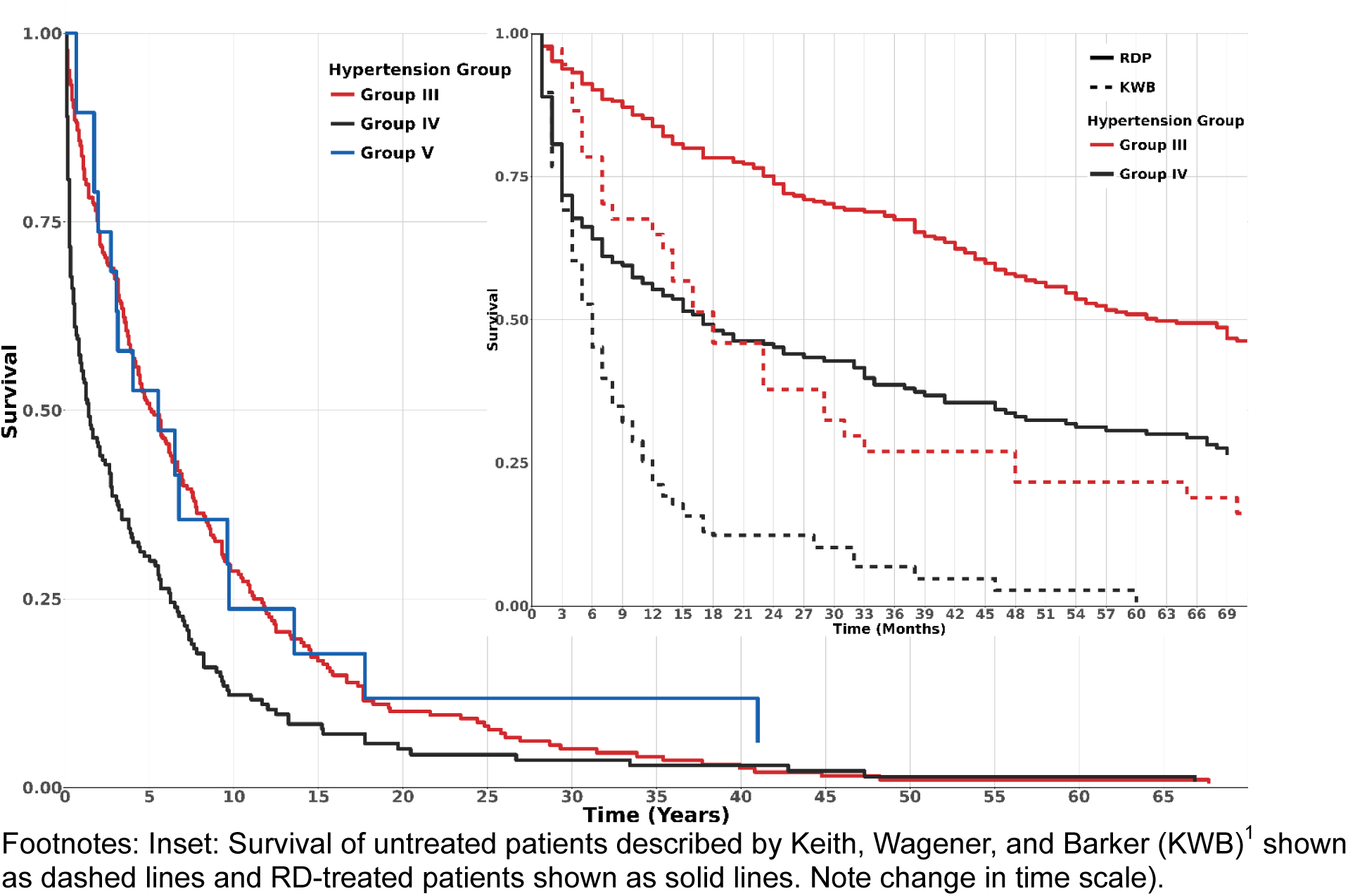
Actuarial survival of MH patients after starting the rice diet.

Of the 454 patients with known dates of death, 19/255 patients in Group III (7.5%); 58/181 patients in Group IV (32.0%), and 0/18 of patients in Group died in ≤90 days. We compared characteristics of patients who died within 90 days with those who died after 120 days (**Table 2**). Those dying within 90 days tended to be younger and male, to have lower initial BMI and greater weight loss over 4 weeks (1.7 vs 0.9 kg/m^2^), to have higher baseline SBP and DBP (225/138 vs 211/127 mmHg) but lesser decrease in BP after 28 days, and to have worse kidney function at baseline (NPN: 67 vs 38 mg/dL; BUN: 41.1 vs 17.6 mg/dL) than those who died after 120 days. Both NPN and BUN increased at week 4, but increases were greater in those who died within 90 days (NPN increase: 13.3 vs 6 mg/dL; BUN increase: 4.3 vs 0.5 mg/dL). Baseline UCl was lower in those who died within 90 days than those who survived >120 days (138 vs 250 mg/dL), but decreased dramatically in both groups to near or better than the goal concentration^3^ of 42 mg/dL by week 4.

**Table 2.**
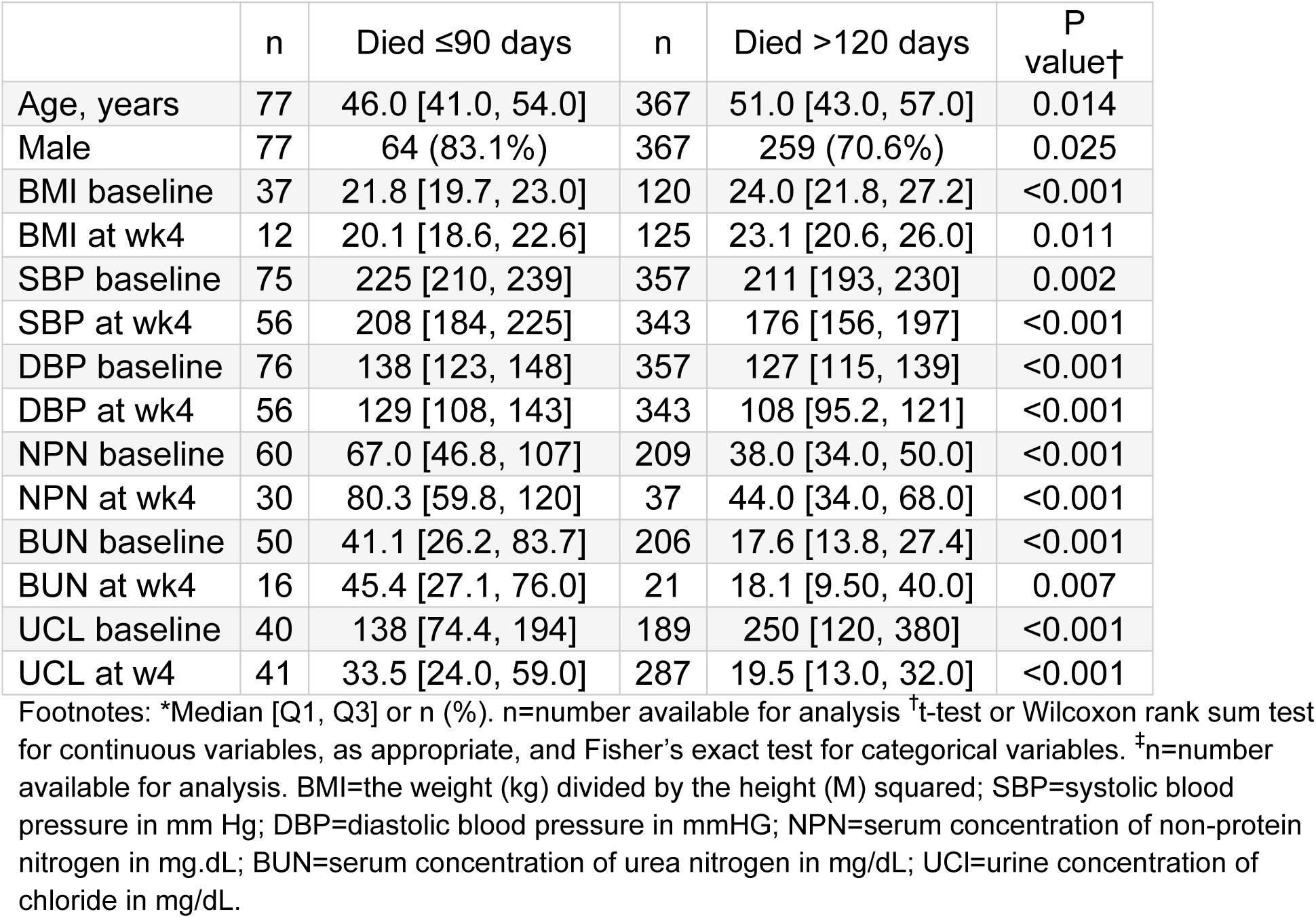
Characteristics of those who died within 90 vs after 120 days*.

|  | n | Died ≤90 days | n | Died >120 days | P value† |
| --- | --- | --- | --- | --- | --- |
| Age, years | 77 | 46.0 [41.0, 54.0] | 367 | 51.0 [43.0, 57.0] | 0.014 |
| Male | 77 | 64 (83.1%) | 367 | 259 (70.6%) | 0.025 |
| BMI baseline | 37 | 21.8 [19.7, 23.0] | 120 | 24.0 [21.8, 27.2] | <0.001 |
| BMI at wk4 | 12 | 20.1 [18.6, 22.6] | 125 | 23.1 [20.6, 26.0] | 0.011 |
| SBP baseline | 75 | 225 [210, 239] | 357 | 211 [193, 230] | 0.002 |
| SBP at wk4 | 56 | 208 [184, 225] | 343 | 176 [156, 197] | <0.001 |
| DBP baseline | 76 | 138 [123, 148] | 357 | 127 [115, 139] | <0.001 |
| DBP at wk4 | 56 | 129 [108, 143] | 343 | 108 [95.2, 121] | <0.001 |
| NPN baseline | 60 | 67.0 [46.8, 107] | 209 | 38.0 [34.0, 50.0] | <0.001 |
| NPN at wk4 | 30 | 80.3 [59.8, 120] | 37 | 44.0 [34.0, 68.0] | <0.001 |
| BUN baseline | 50 | 41.1 [26.2, 83.7] | 206 | 17.6 [13.8, 27.4] | <0.001 |
| BUN at wk4 | 16 | 45.4 [27.1, 76.0] | 21 | 18.1 [9.50, 40.0] | 0.007 |
| UCL baseline | 40 | 138 [74.4, 194] | 189 | 250 [120, 380] | <0.001 |
| UCL at w4 | 41 | 33.5 [24.0, 59.0] | 287 | 19.5 [13.0, 32.0] | <0.001 |

Footnotes: \*Median [Q1, Q3] or n (%). n=number available for analysis †t-test or Wilcoxon rank sum test for continuous variables, as appropriate, and Fisher's exact test for categorical variables. ‡n=number available for analysis. BMI=the weight (kg) divided by the height (M) squared; SBP=systolic blood pressure in mm Hg; DBP=diastolic blood pressure in mmHG; NPN=serum concentration of non-protein nitrogen in mg.dL; BUN=serum concentration of urea nitrogen in mg/dL; UCL=urine concentration of chloride in mg/dL.

Figure 3 shows median values of interval-average SBP and DBP in patients who died during their first ten years after starting the RD compared to those who lived 10 years or more. SBP was significantly different between groups at baseline and over the ensuing 6 years; DBP was significantly different at baseline through year 3.

**Figure 3.**
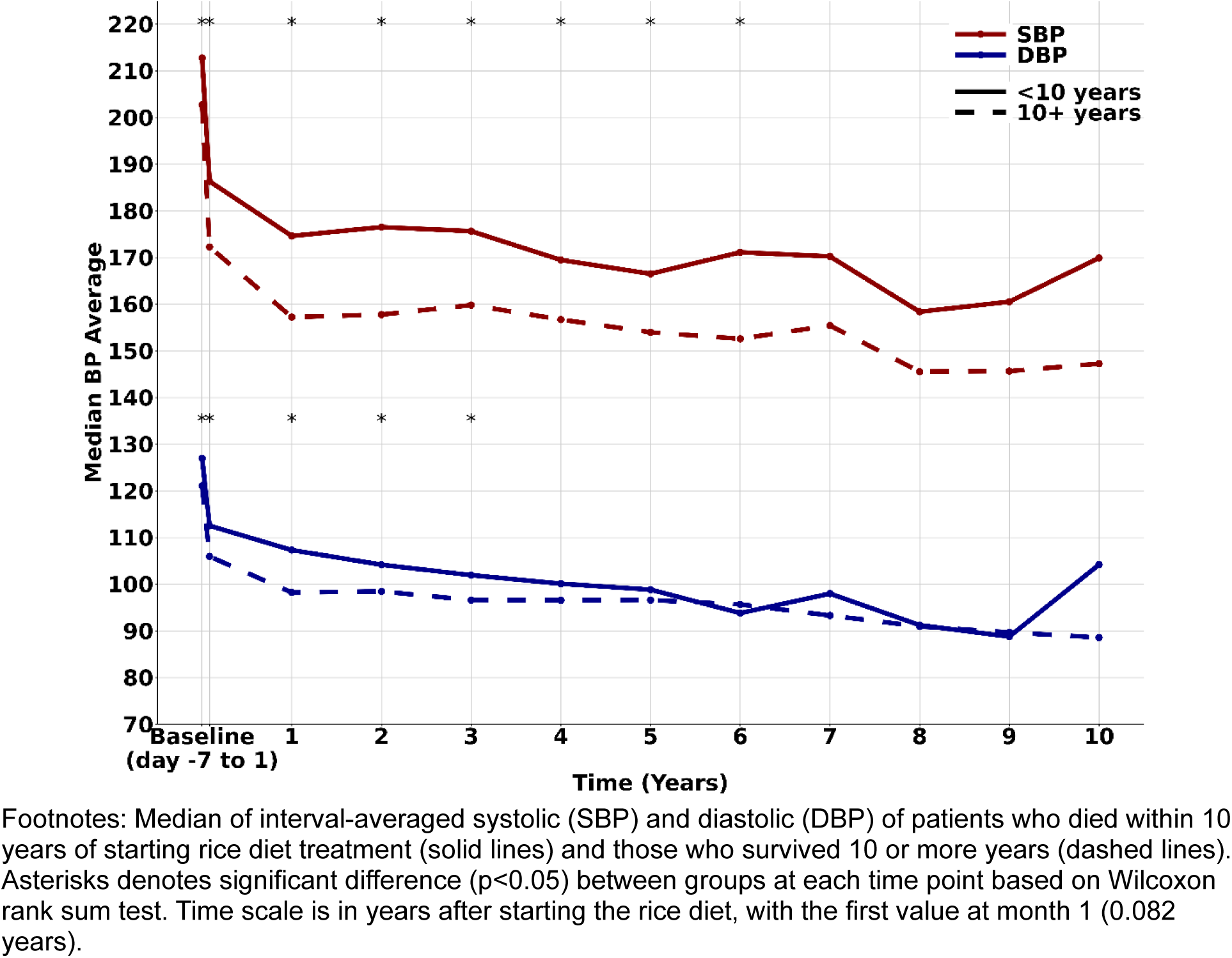
Blood pressure in patients who survived less than or more than 10 years.

Figure 4 presents actuarial probability estimates of time to resolution of retinal H and P over one year after starting the RD. The two ocular findings, H and P, were analyzed without respect to hypertension groups (i.e., Groups III and IV were combined for analysis of H, as were Groups IV and V for analysis of P). Over the course of one year after starting the RD, retinal H resolved well, with 50% resolution by 180 days (or less), and nearing 90% at 360 days (lessening of H was sometimes noted but we considered only complete clearance as resolution). P cleared in ∼50% of patients by 120 days (or less) and in 107 of the 110 (out of an initial 143) patients who survived one year.

**Figure 4.**
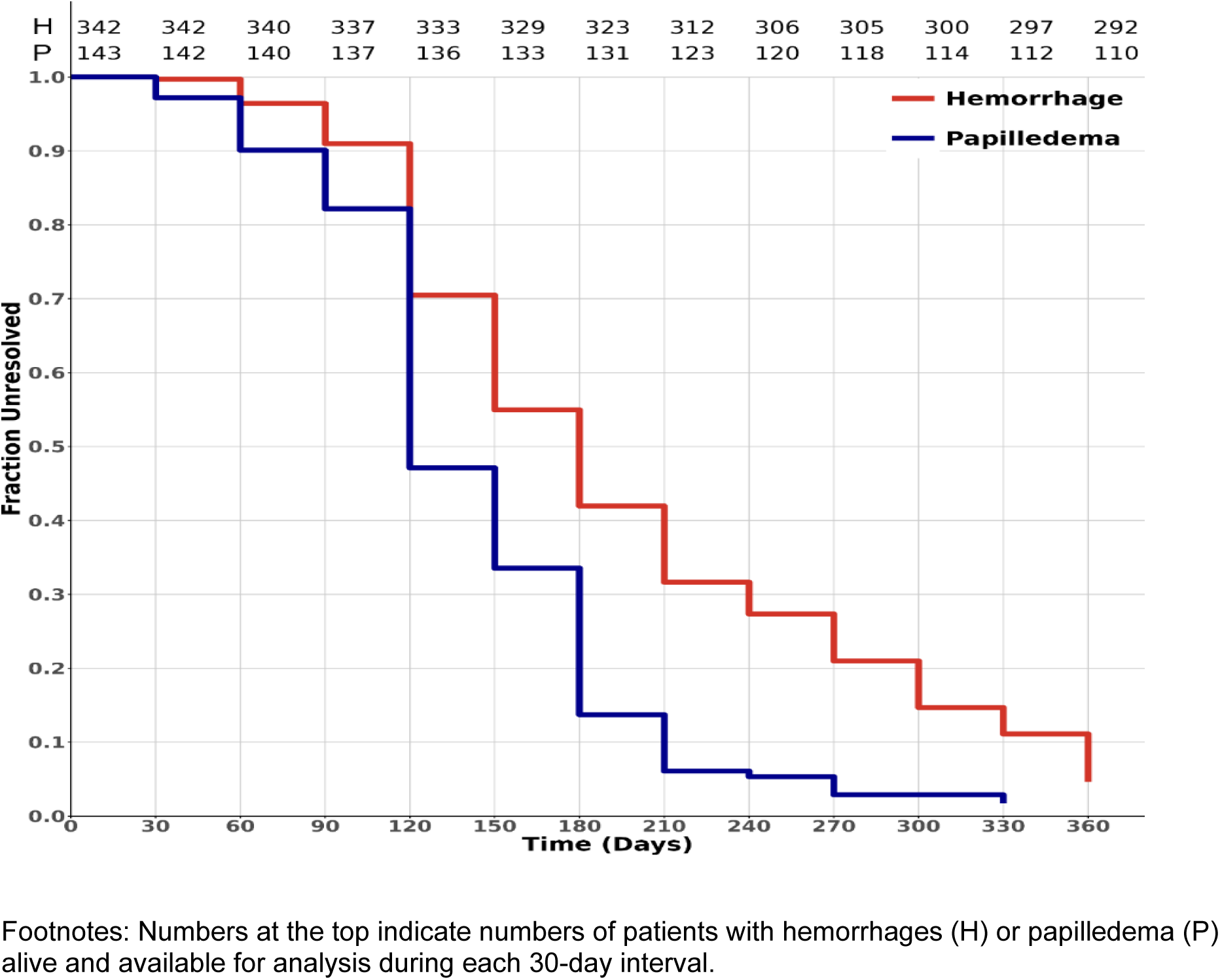
Actuarial estimates of resolution of retinal hemorrhage and papilledema.

## DISCUSSION

In this cohort of 544 patients, those in Group III tended to be older than those in Group IV or V (**Table 1**), but BMI and male predominance were similar across groups (possibly greater in Group IV). The three hypertension groups showed significant differences in BP and kidney function both at baseline and after 4 weeks on the RD. Patients in Group IV appear “sicker” than those in Group III: baseline SBP, DBP and NPN were higher, and the BP-lowering at one month was less in Group IV than III; the impression of greater illness burden is consistent with differences in duration of life after starting the RD: 1.0 years for those in Group IV vs 4.2 years in Group III (supplemental **Table S4**). Group V patients appear to lie between Groups III and IV, being similar to Group III in baseline, one-month SBP response, actuarial survival estimate, and duration of life (3.6 years, **Table S4**), but more like Group IV patients in baseline and one-month DBP response. The small number of patients in Group V prohibits finer distinction.

Serum levels of non-protein nitrogen (NPN) and urea nitrogen (BUN) provide the best available (although now outmoded) indicators of renal status in these patients.

Data in **Table 1** imply that kidney function was modestly impaired at baseline in all groups, but worse in Group IV than Group III; after 4 weeks of RD treatment, NPN and BUN decreased slightly in Group III, but increased in Groups IV and V, suggesting further deterioration of kidney function. Since blood levels of NPN and BUN fall with reduced protein intake, and may increase transiently after a rapid decrease in renal artery perfusion pressure, it is not certain that kidney function improved in Group III, but it is likely that it deteriorated, despite RD treatment, in Group IV, and possibly, Group V.

### Long-term Blood Pressure Effects of Dietary Treatment

As shown in Figure 1, SBP and DBP decreased substantially over the first 1-2 years after enrollment in the RD, with >30% of the overall reduction in excess BP occurring during the first month of RD (Figure 1, Panel C). Both SBP and DBP declined slowly over years 1-9, becoming relatively stable thereafter at a median of ∼143/87 mmHg. The pattern shown in Figure 1 does not appear to be impacted by selective loss of patients with the highest BPs; comparison of median BPs in the entire cohort with those of patients who survived 120 days or more shows only trivial differences (1-2%; supplementary **Table S2**) and no change in temporal distribution. As effective antihypertensive medications became available after 1958, the proportion of RD patients being treated rose to about half (Figure 1). However, we are confident that the initial BP response and effects on mortality can be ascribed to the RD because so few patients (48/544; 8.8%) were treated with even feebly effective anti-hypertensive medication during their crucial first month of RD treatment.^3^

The relatively infrequent use of medications by long-term survivors may reflect prevailing attitudes about the “essentiality” of hypertension, or contemporaneous satisfaction with SBP in the range of 140-160 mmHg. Despite the increasing availability of suitable anti-hypertensives over time, that availability did not seem to affect survival; there was minimal to no correlation (Pearson’s r = 0.044) between duration of survival and calendar date of starting the RD (supplemental **Figure S1**).

### Survival with Rice Diet

Patients in Group IV had shorter survival (median of ∼510 days) than those in Group III (∼1890 days), but both groups survived substantially longer than the similar, untreated patients described by KWB^1^ (median survivals: ∼180 for Group IV and ∼540 days for Group V). It seems reasonable to ascribe the differences to the dietary treatment since no effective anti-hypertensive drugs were available to KWB patients and only 8.8% of RD patients were treated (at least, initially) with any (mostly ineffective) antihypertensive medications at all.

Patients with Group IV hypertension had a shorter median life expectancy than those with Group III hypertension, largely because 32% of patients in Group IV died during the first 90 days on the RD (Figure 2 Inset). After 90-120 days, survival curves for Group III and IV RD patients diverge from those described by KWB. One year after RD treatment, and increasingly so thereafter, survival curves for RD Group III and Group IV become almost indistinguishable, so if Group IV patients can live long enough, their survival essentially becomes that of Groups III and V.

Why the therapeutic response to the RD takes months to become apparent is not immediately obvious. However, data in **Table 2** imply that those who died early, although younger, were “sicker” in terms of BP elevation, impaired BP-lowering response to the diet, worse baseline kidney function, and lower BMI, suggesting malnutrition, than those who died later. In addition, UCl was lower at baseline in those who died early, possibly further reflecting poor food intake and malnutrition at entry. It may well be that patients who die in the first few months have already passed a point-of-no-return and cannot survive long enough to reap the benefits of the dietary intervention. Modern renal replacement therapy may help by providing time for healing,^9^ an option not available for most of Kempner’s patients.

Initial severity, as well as magnitude and rapidity of early BP response to RD treatment, appear related to survival. Figure 3 shows that patients who died within 10 years after starting the RD had higher initial BP and somewhat less BP lowering compared to those who lived 10 years or more. During the first month and first year of RD treatment, SBP in the group that survived <10 years reached ∼ 25% and ∼60% of the overall lowering achieved, while the group that survived ≥10 years reached ∼45% and ∼75%, respectively (DBP changes were similar).

Calculation of Cox proportional hazards ratios (HR) for mortality by age; sex; initial BMI; baseline values for SBP, NPN, BUN, UCl, and the magnitude of BP decline over the first 28 days of RD treatment shows similar results (supplementary **Table S3**).

HR for long-term mortality decreases dramatically according to the magnitude of SBP reduction during the first month. Other factors that affect survival include male sex (compared to female, HR for long-term mortality was 1.5; p <.005), and lesser but still significant HRs for higher baseline SBP and NPN, the latter supporting the conclusion of Newborg and Kempner^2^ that worse kidney function increases mortality.

Not clear from our observational data is what prompts a better or worse BP response to the RD. However, the implication that intense and more BP-lowering is associated with a survival benefit would argue in favor of using now-available effective anti-hypertensive medications to attain faster and further reduction in BP, but we should not lose sight of the value of intensive dietary intervention.

### Resolution of Ocular Abnormalities

Treatment with the RD allowed resolution of the retinal H and P that define MH. Figure 4 indicates half-times to resolution of ≤180 days for H and ≤120 days for P; after one year, H had cleared in 235/293 patients (out of an original 342) who survived one year, and P in 107/111 (out of an original 143 patients) who survived one year. As with overall survival, rates of resolution of retinal H and P appear to increase after ∼100 days of RD therapy, but because retinal photographs were taken at non-standard (almost haphazard) intervals, resolution might, in fact, have occurred weeks or months before the conclusive negative examination. Our data, therefore, indicate that the longest possible time until resolution. Nevertheless, median time to resolution was no greater, and possibly less than 120-180 days, and P appears to resolve more quickly than H.

### Persistent Loss of life Expectancy

RD treatment improved life expectancy but did not restore it to that anticipated for U.S. population at large. A severe toll was extracted in terms of years of potential life lost, ranging from 12.3 years for men with Group III hypertension to 25.7 years for women with Group IV hypertension (supplemental **Table S4**). Median survival of patients with Group IV hypertension was less than for Groups III and V, largely because of the early mortality in Group IV (Figure 2 Inset); when Group IV patients lived >120 days, their survival approximated that of Group III patients (supplemental **Table S4**). Loss of potential life is further demonstrated by the fact that only 34/454 (7.5%) patients lived beyond the average expected for their age and birth-year. The calculated toll in years of potential life lost appears greater for Group V (20.3 vs 12.7 for Group III and 20.1 for Group IV) mainly because the small Group V cohort was younger when they started (42.0 years vs 54.0 for Group III and 46.8 for Group IV) and therefore had more years of potential life to lose.

One question surrounding MH is whether the occurrence of P indicates a more lethal evolution of MH than retinal H alone. Lip et al^5^ concluded that P without H conferred a worse prognosis than P with H. However, as shown in Figure 2, the survival curve for patients in our Group V (P without H) closely tracked that of Group III (H without P), but not IV (P with H). Similarly, Cox proportional hazards analysis gave a HR for mortality (data not shown) of 1.77 (p <.001) for Group IV vs Group III, but only 1.10 for Group V vs Group III (p=0.686). These observations suggest that MH does not progress in severity through defined stages (i.e., high BP, then H, then P), but rather that these features provide separate but not sequential risks to survival. Overall risk would be determined additively by the number of organ systems affected (arterial pressure, retina, brain, kidney, etc.). If a breakdown of vascular autoregulation underlies MH, different organs may have differing autoregulatory thresholds. Thus, the involvement of any one is bad, two is worse, and three, even worse.

For Kempner, as today, MH was a medical emergency, but high-income countries now see few such patients, and the MH syndrome has largely disappeared from Kempner’s hospital.^3^ Current thoughts on pathogenesis, the possible role of inflammation, immunological factors, and thrombotic microangiopathy have been reviewed by Boulestreau et al,^10^ but it is not clear why this frightening syndrome has faded from view. We rather doubt that widespread present availability of pharmacotherapy or medical advances alone explain the decrease in prevalence because so many individuals are unaware of, or have uncontrolled, severe hypertension.^11^ Alterations in dietary intake do not explain the phenomenon: sodium intake has not, to our knowledge, decreased, potassium intake has surely not increased, and calorie intake has risen over time. Perhaps because so few non-ophthalmologists examine optic fundi, today’s MH patients are classified as having “hypertensive emergency” or “posterior reversible encephalopathy syndrome” rather than MH.

### Strengths and Limitations

Strengths of this report include: 1) A large cohort of patients with MH, more than 90% of whom were treated, at least initially and during their first months, only with diet rather than antihypertensive drugs. 2) Treatment at a single, dedicated center, with compliance to the ultra-low sodium diet enhanced by provision of food and monitored by urine chloride testing. 3) Frequent monitoring of BP. 4) Long follow up. 5) Availability of data about resolution of ocular fundus abnormalities, and long-term survival.

There are also several limitations: 1) Although the database available for analysis was constructed from carefully preserved and extensive records of clinical care, it was not driven by a well-defined experimental protocol. As a result, the data are not uniform in time or extent of collection. 2) Because all treatment occurred in Durham, NC, many patients were only temporarily in residence there, returning to their sometimes distant residences for further care. As a result, continuous observation and monitoring was interrupted. 3) Lack of documentation of date of death for a portion (17%) and lack of cause of death for almost all patients is a detriment. 4) Irregular and infrequent monitoring of ocular fundus changes prohibits precise identification of time to resolution of those changes. 5) The unavailability of modern markers of kidney function and lack of systematic use of available measures such as serum NPN is a detriment.

### Conclusions

The salutary effects conferred by the RD (prolonged survival and resolution of ocular fundus abnormalities) most likely reflect temporal improvement (healing) of the multi-organ arteriolar disease^12^ or whatever else may underlie MH. Our data confirm the beneficial effect of the RD in treating MH, without disputing the benefit of antihypertensive drug therapy,^9^ but emphasizing that strict control of diet, especially sodium intake, can be a useful adjunct in treating even the most severe forms of hypertension. Despite the concerns of some that low sodium diets may be deleterious,^13^ our data show a distinct survival benefit (and no discernible ill effects) from long-term prescription of the lowest conceivable intake of dietary sodium. The RD was also extremely low in fat, protein and chloride content, but the careful experimental observations of Dole et al^14^ indicate that sodium deprivation is likely the main driver of the survival benefit seen in Figure 1. We see this report as evidence that food can, indeed, serve “as medicine.”

### Perspectives

The ultra-low sodium rice diet (RD) lowered the high blood pressure (BP) of malignant hypertension (MH), but little is known about how it affected overall survival or resolution of the ocular hemorrhages and papilledema that define MH. We analyzed those outcomes in 544 patients with MH treated with RD between 1942-1982. Compared to untreated patients with MH, RD treatment tripled median survival in those with retinal hemorrhages (Group III) and those with both retinal hemorrhages and papilledema (Group IV). Survival of patients with papilledema alone (Group V) paralleled that of Group III, rather than Group IV. Antihypertensive drug therapy played little role; such medications were used in <10% of patients during the first month of RD and in only about half of long-term survivors.

BP response after 4 weeks on RD correlated with lower HR for mortality, but despite clear benefits, the RD did not restore longevity to that of the population at large; experiencing MH inflicted a loss 12.4-25.7 years of anticipated life. Ocular hemorrhages and papilledema resolved on the RD; median time to resolution was ≤180 days; hemorrhages resolved in 90 % of patients who survived ≤1 year, and P in almost all.

The salutary effects of RD demonstrate the value of true restriction of sodium, at least as an adjunct, in the treatment of hypertension.

## Data Availability

Thee data files used in this manuscript will be posted in a Duke repository at time of publication.

## Acknowledgments

FAN, PJK and PHL contributed to the concept of the manuscript. FAN drafted the first version of the manuscript. JOL, FAN and YJL analyzed the data. AB and WM organized and provided data for analyses. All authors provided critical input to analyses, interpretation of results, review and edits. PHL is the guarantor of the manuscript. The authors gratefully acknowledge the late Dr. E. Harvey Estes who catalyzed the formation of the team that carried out this project.

## List of nonstandard abbreviations and acronyms

BP: blood pressure
BUN: blood urea nitrogen
DBP: diastolic blood pressure
IQR: Interquartile Range
KWB: Keith, Wagener, Barker
MH: malignant hypertension
NPN: non-protein nitrogen
RD: rice diet
SBP: systolic blood pressure
UCl: urine chloride concentration

## Pathophysiological Novelty and Relevance

### · What is new?

Although dietary treatment lowers blood pressure, it has not hitherto been known whether such treatment affects long-term survival or the clearance of the ocular hemorrhages and papilledema that define malignant hypertension (MH). The present results show that the rice diet, which is extremely low in sodium, fat and protein, provided a survival benefit and led to clearing of retinal hemorrhage and papilledema. The findings buttress the argument in favor of food as medicine.

### What is Relevant?

Our data, showing prolonged survival and ocular improvement, promote the role of dietary intervention in the treatment of hypertension, especially severe or malignant hypertension.

### Clinical/Pathophysiological Implications?

Restricted intake of sodium (and possibly of fat and protein) lowers blood pressure and improves survival and other abnormalities of severe hypertension, but by itself, may not eliminate all deleterious effects. This argues in favor of effective pharmacological treatment but also indicates a relevant and beneficial role for intense diet therapy. Further evaluation might elucidate the mechanism of the hypotensive effects and determine the optimal level of sodium restriction that will maximize benefits.

## References

1. Keith NM, Wagener WH, Barker NW. Some different types of essential hypertension: Their course and prognosis. The American journal of Medical Sciences. 1939;197:332–343.

2. Newborg B, Kempner W. Analysis of 177 cases of hypertensive vascular disease with papilledema. Am J Med. 1955;19:33–37.

3. Sanoff SL, Klemmer PJ, Neelon FA, La, JO, Lopez D, Bohannon A, McDowell WK, Luft FC, Li Y-J, Lin P-H. Treating malignant hypertension with a low-sodium, low-protein, low-fat diet. Hypertension. 2026;83:26–36.

4. Sommerfeld R, Ermler P, Fehr J, Bergner B, Lopez D, Sanoff S, Neelon FA, Kuo A, McDowell W, Li Y-J, Fox S, Ghajar A, Gensch E, Lorenz C, Preiss M, Richter T, Luft FC, Klemmer P, Bohannon A, Lippert C, Lin P-W. Modern perspective of the Rice Diet for hypertension and other metabolic diseases. BMJ Nutrition, Prevention & Health 2024;7:e000949. doi:10.1136/bmjnph-2024-000949.

5. Lip GYH, Beevers M, Dodson PM, Beevers DG. (1995). Severe hypertension with lone bilateral papilloedema: a variant of malignant hypertension. Blood Press. 1995;4(6):339–342.

6. Social Security Administration.ssa.gov/OACT/Downloadables/CY/index.html. Accessed June 19, 2025

7. Parkin DM, Hakulinen T. Analysis of survival. Chapter 12 in Jensen OM, Parkin DM, MacLennan R, Muir CS, Skeet R (ed). Cancer Registration: Principles and Methods Oxford University Press, 1991.

8. Cox DR. Regression models and life-tables. J R Stat Soc Ser B Methodol. 1972;34:187–220

9. Woods JW, Blythe WB. Management of malignant hypertension complicated by renal insufficiency. N Engl J Med. 1967;277:57–61.

10. Boulestreau R, Spiewak M, Januszewicz A, Kreutz R, Guzik TJ, Januszewicz M, Muiesan ML, Persu A, Sarafidis P, Volpe M, Zaleska-Zmijewska A, van den Born B-JH, Messerli FH. malignant hypertension: a systemic cardiovascular disease. J Am Coll Cardiol 2024;83:1688–1701.

11. Richardson LC, Vaughan AS, Wright JS, Coronado F. Examining the hypertension control cascade in adults with uncontrolled hypertension in the US. JAMA Network Open. 2024;7(9):e2431997. doi:10.1001/jamanetworkopen.2024.31997.

12. Wagener HP, Keith NM. Diffuse arteriolar disease with hypertension and the associated retinal lesions. Medicine,1939;18(3):317–430.

13. Graudal N, Jurgens G, Baslund B, Alderman MH. compared with usual sodium intake, low- and excessive sodium diets are associated with increased mortality: a meta-analysis. Am. J. Hypertens. 2014;27(9):1129–1137.

14. Dole VP, Dahl LK, Cotzias GC, Eder HA, Krebs ME. Dietary treatment of hypertension. Clinical and metabolic studies of patients on the rice-fruit diet. J Clin Invest. 1950;29(9):1189–1206.

